# Three Decades of FDA Authorizations of AI/ML-Enabled Medical Devices: Persistent Specialty Concentration and the Care-Delivery Gap (1995–2025)

**DOI:** 10.64898/2026.05.08.26352766

**Authors:** Pouyan Golshani, Mary Joseph

## Abstract

The US Food and Drug Administration (FDA) maintains a public list of artificial intelligence and machine learning (AI/ML)-enabled medical devices that have received marketing authorization. Prior published analyses examined this list at earlier time points and reported a marked dominance of radiology applications. We performed a cross-sectional analysis of all 1,430 AI/ML-enabled medical device authorizations recorded by the FDA between September 1995 and December 2025 to characterize the cumulative growth, specialty distribution, and manufacturer concentration of authorized devices. The annual authorization volume increased from a mean of 1.8 per year between 1995 and 2014 to 264 per year between 2023 and 2025, with 331 authorizations recorded in 2025 alone. Devices reviewed by the FDA’s Radiology panel accounted for 1,094 of 1,430 authorizations (76.5%), and the three most represented panels (Radiology, Cardiovascular, and Neurology) accounted for 90.6% of all authorizations. Several large clinical specialties were represented by very small numbers of authorized devices, including Pathology (n = 9), Microbiology (n = 6), and Obstetrics and Gynecology (n = 4). No authorizations were recorded under a psychiatry or behavioral health review panel. Of 740 unique companies, 502 (67.8%) had a single authorized device, while 13 companies (1.8%) accounted for 217 devices (15.2%). The cumulative regulatory record demonstrates rapid growth that has been concentrated in image-rich diagnostic specialties, with limited representation across many specialties that account for substantial clinical activity in the United States. These findings may inform policy discussions about where regulatory, infrastructure, and dataset investments are most needed to broaden the clinical scope of medical AI.

## Introduction

The number of artificial intelligence and machine learning (AI/ML)-enabled medical devices that have received marketing authorization from the US Food and Drug Administration (FDA) has grown rapidly over the past decade [1]. The FDA maintains a public list of these devices, which it updates periodically and which has been the basis of multiple published analyses characterizing the regulatory landscape [1,2,3]. An early review identified 64 AI/ML-based medical devices through 2020 [1]. A subsequent landscape analysis using the same FDA list identified 691 devices by October 2023 [2]. A more recent taxonomic review examined 1,016 authorizations to characterize the underlying AI tasks performed by these devices [3]. Across these analyses, devices reviewed by the FDA’s Radiology panel have consistently represented the majority of authorizations [1,2,3].

Despite the cumulative growth of the regulatory record, several gaps in published analyses remain. First, no published analysis has characterized the full cumulative dataset through the end of calendar year 2025, a period during which authorization volume continued to accelerate. Second, prior analyses have generally reported the radiology share without quantifying the corresponding underrepresentation of other clinical specialties relative to their burden of clinical activity. Third, the concentration of authorizations among a small number of repeat-authorizing manufacturers has not been systematically described, despite implications for market structure and innovation incentives.

In this report, we describe the cumulative regulatory record of AI/ML-enabled medical devices authorized by the FDA between September 1995 and December 2025. We characterize the growth trajectory by year, the distribution of authorizations across the FDA’s lead review panels, the temporal stability of specialty concentration, and the distribution of authorizations across manufacturers.

## Materials and Methods

### Data source

This analysis used the FDA’s publicly maintained list of AI/ML-enabled medical devices, accessed in a deidentified tabular form [4]. The FDA-maintained list reports devices that have received marketing authorization through the 510(k), De Novo, or Premarket Approval (PMA) pathways and that the FDA has identified as containing AI/ML functionality [4]. The dataset contained 1,430 authorizations with final-decision dates between September 29, 1995, and December 30, 2025. The same regulatory record is also accessible through PhysicianAITools.com (https://physicianaitools.com), a publicly available aggregator catalog that integrates the FDA list with additional discovery-based metadata not analyzed in the present study; analyses reported here used the FDA’s source list directly.

### Variables

For each authorization, the dataset contained the date of final decision, submission number, device name, manufacturer (company), the FDA lead review panel (an FDA classification corresponding broadly to the medical specialty of intended use), and the primary product code. Submission numbers were used to classify authorizations by regulatory pathway: numbers beginning with “K” were classified as 510(k); numbers beginning with “DEN” as De Novo; and numbers beginning with “P” as PMA.

### Analysis

Descriptive analyses were performed using Python 3.11 with the pandas (version 2.x) library. We computed annual and cumulative authorization counts by year of final decision; the proportion of authorizations attributable to each FDA lead review panel; and the distribution of authorized devices across unique manufacturers. To examine the temporal stability of specialty concentration, we computed the proportion of authorizations attributable to the Radiology panel and to the top three combined panels for each year between 2015 and 2025. The earlier period (1995–2014) was excluded from year-stratified specialty analysis because of small annual counts. No formal hypothesis tests were planned; analyses were descriptive.

### Ethics

This analysis used a publicly available regulatory dataset that contained no patient-level information. Institutional review board approval was therefore not required.

## Results

### Cumulative growth, 1995–2025

A total of 1,430 AI/ML-enabled medical device authorizations were recorded between September 29, 1995, and December 30, 2025. The mean annual authorization rate was 1.8 per year between 1995 and 2014, increasing to 39.2 per year between 2015 and 2019, 135.3 per year between 2020 and 2022, and 264.0 per year between 2023 and 2025. The annual authorization count reached a single-year maximum of 331 authorizations in 2025. The cumulative count crossed 100 authorizations in 2018, 500 in 2022, and 1,000 in 2024 (Figure 1).

**Figure 1.**
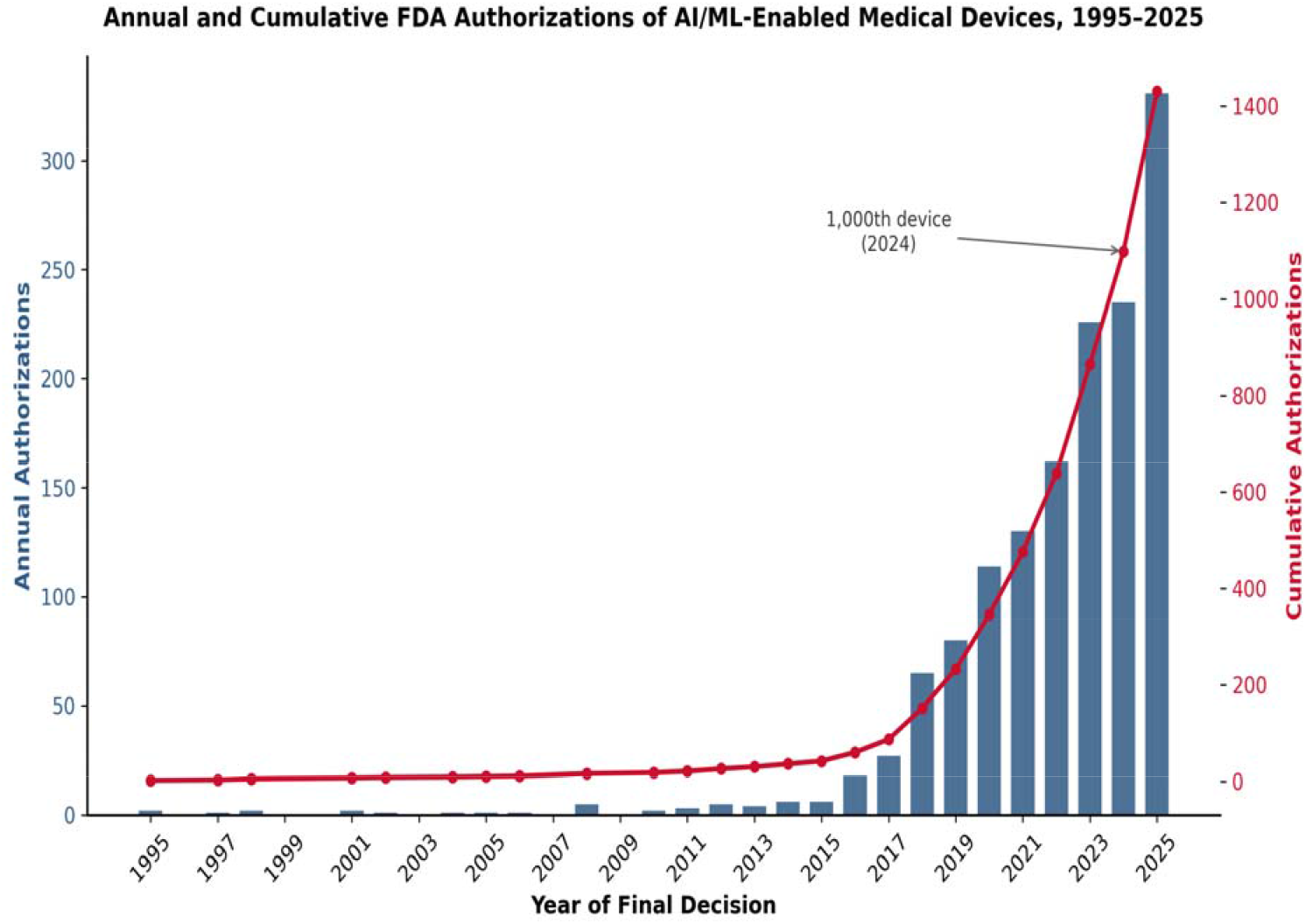
Annual and cumulative FDA authorizations of AI/ML-enabled medical devices, 1995–2025.

### Specialty distribution

Authorizations were unevenly distributed across the FDA’s lead review panels. The Radiology panel accounted for 1,094 of 1,430 cumulative authorizations (76.5%), the Cardiovascular panel for 136 (9.5%), and the Neurology panel for 65 (4.5%). The three most represented panels collectively accounted for 1,295 authorizations (90.6%), and the five most represented panels accounted for 1,345 authorizations (94.1%) (Figure 2).

**Figure 2.**
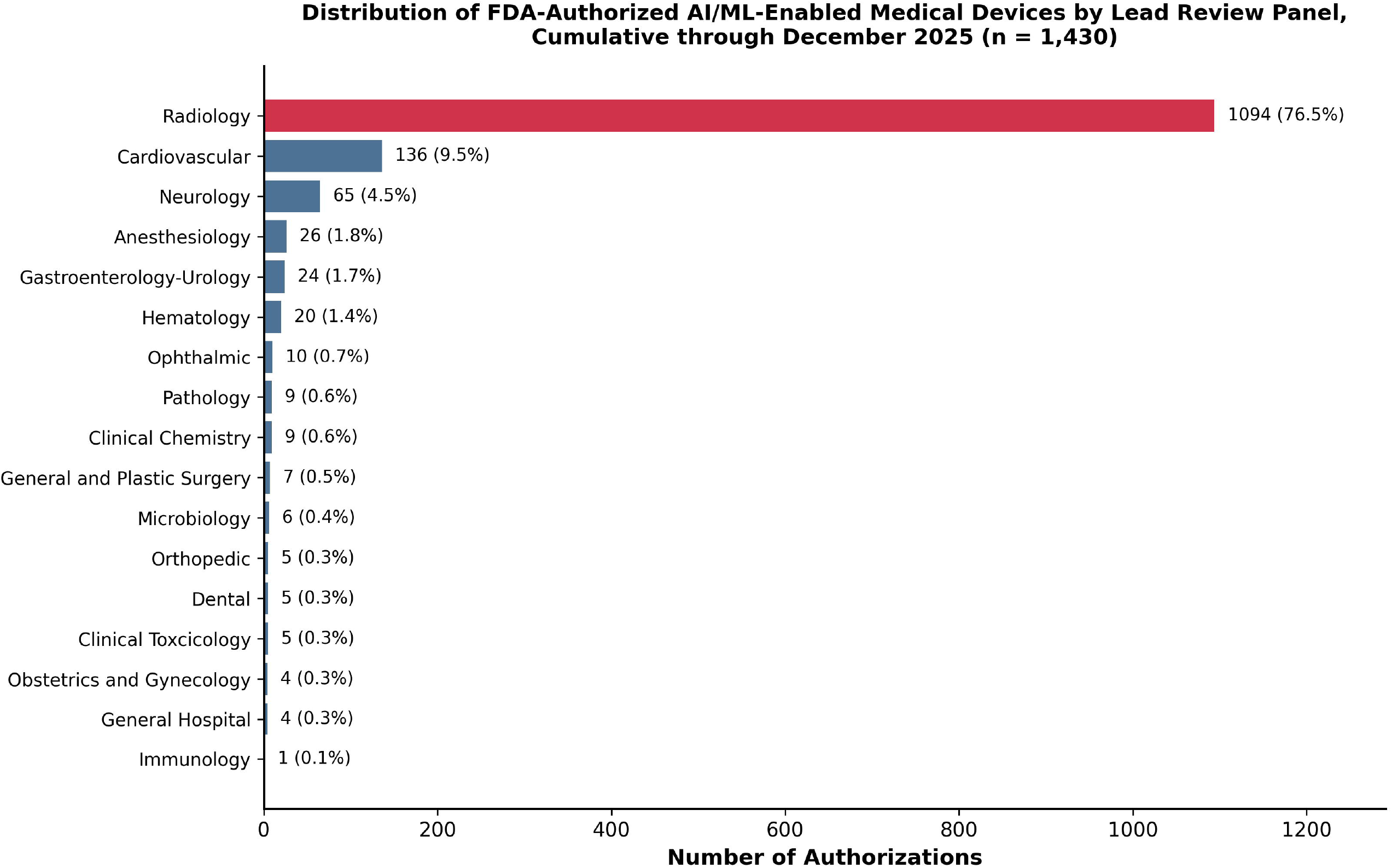
Cumulative distribution of FDA-authorized AI/ML-enabled medical devices by lead review panel, 1995– 2025 (n = 1,430).

Several panels corresponding to large clinical specialties were associated with small absolute numbers of authorizations. The Pathology panel was associated with 9 authorizations (0.6%), the Microbiology panel with 6 (0.4%), the Dental panel with 5 (0.3%), and the Obstetrics and Gynecology panel with 4 (0.3%). No authorizations in the dataset were classified under a psychiatry or behavioral health review panel.

The radiology share of annual authorizations was stable over time. Between 2018 and 2025, the proportion of annual authorizations reviewed by the Radiology panel remained between 61.5% and 86.8%, with no sustained downward trend across the most recent decade (Figure 3). In each of the years 2020 through 2025, the Radiology panel accounted for at least 76% of new authorizations.

**Figure 3.**
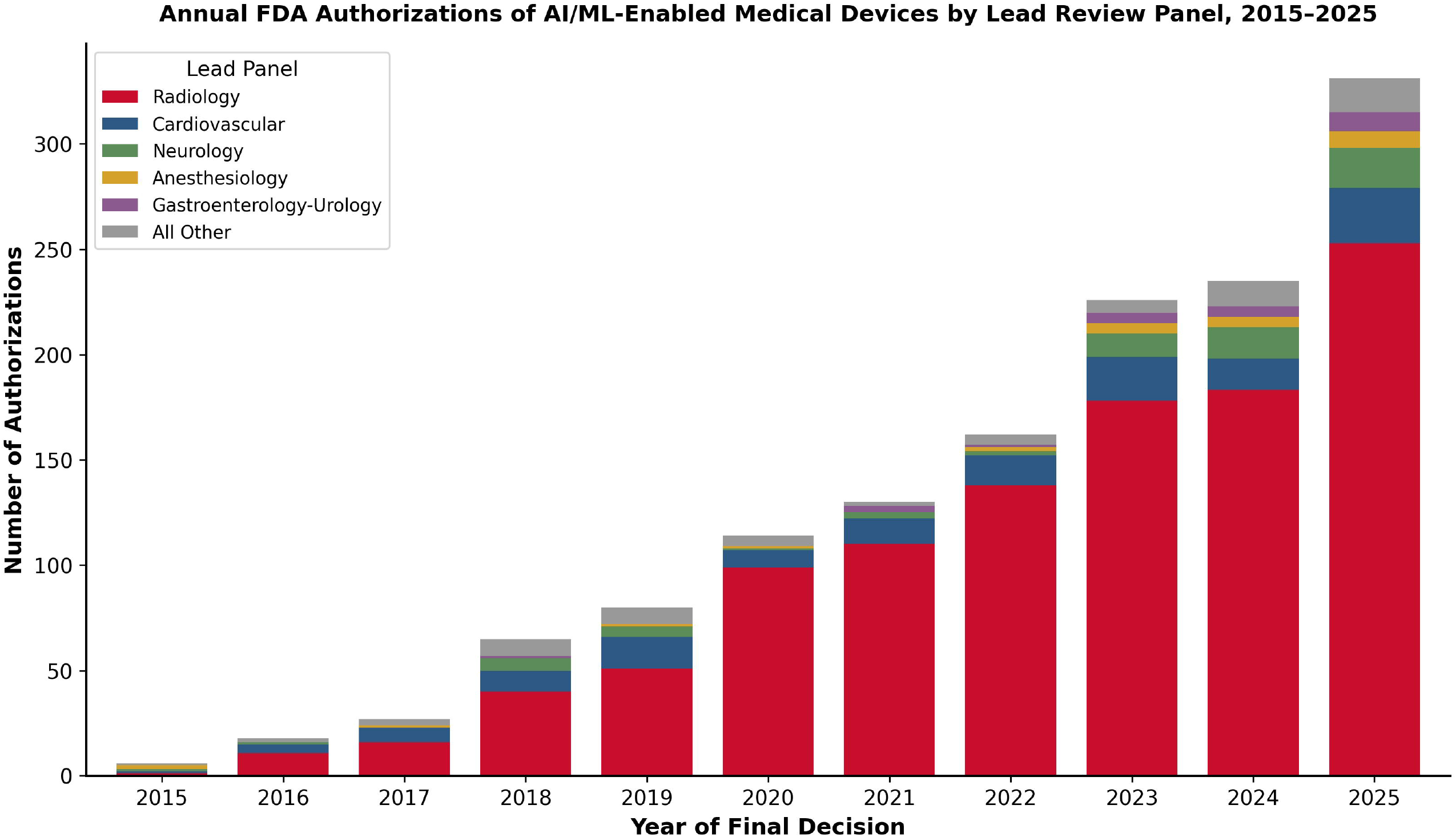
Annual FDA authorizations of AI/ML-enabled medical devices by lead review panel, 2015–2025.

### Regulatory pathway

Of 1,430 authorizations, 1,376 (96.2%) were cleared through the 510(k) pathway, 37 (2.6%) through the De Novo pathway, and 17 (1.2%) through the PMA pathway. The 510(k) share is consistent with prior reports identifying 510(k) clearance as the predominant authorization mechanism for AI/ML-enabled devices, including a recent analysis of the 2024 cohort that reported 94.6% of that year’s AI/ML authorizations cleared via 510(k) [2,3,6].

### Manufacturer concentration

The 1,430 authorizations were associated with 740 unique manufacturers. Of these, 502 manufacturers (67.8%) had a single authorization in the dataset. A total of 13 manufacturers (1.8%) had 10 or more authorizations and collectively accounted for 217 of 1,430 authorizations (15.2%). The 10 manufacturers with the highest authorization counts accounted for 217 authorizations (15.2%), and the 25 manufacturers with the highest counts accounted for 336 authorizations (23.5%) (Figure 4). The single manufacturer with the highest authorization count had 51 authorizations.

**Figure 4.**
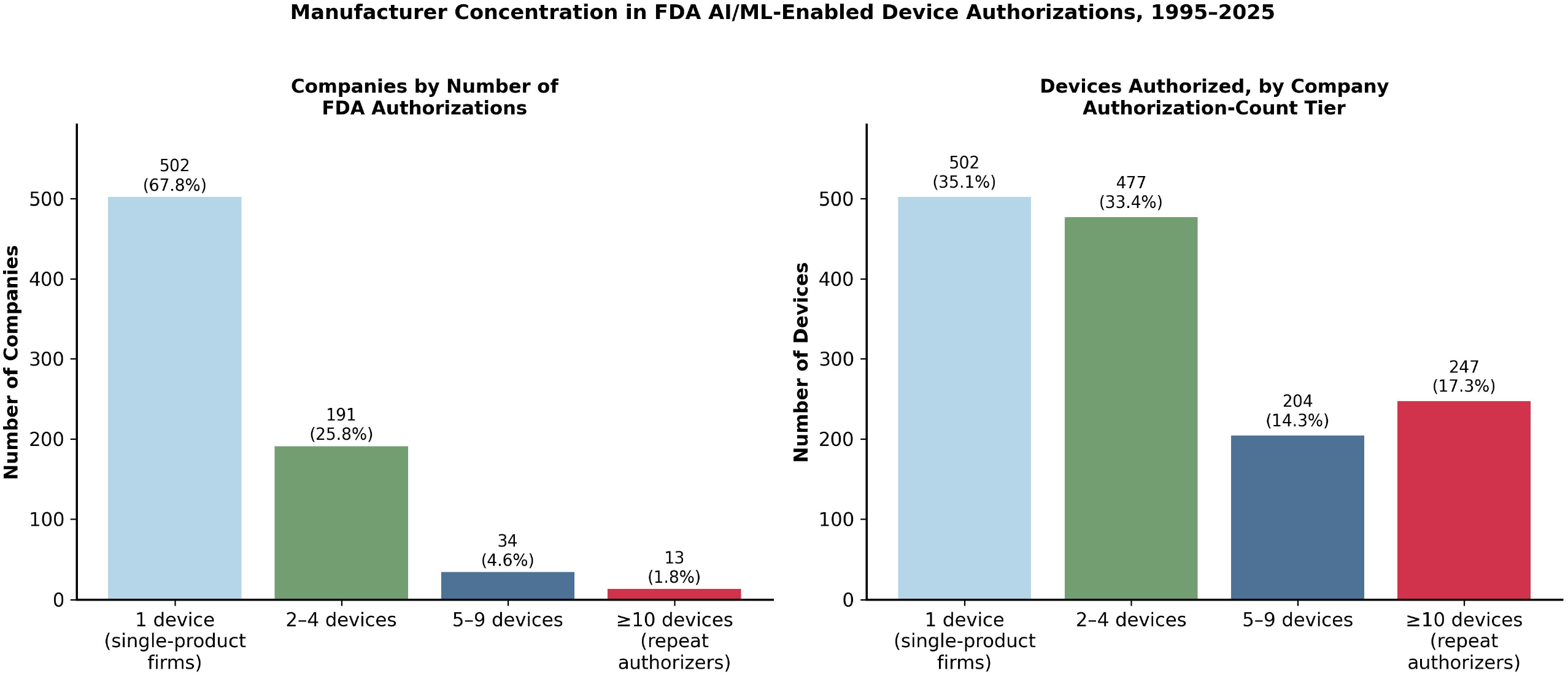
Manufacturer concentration in FDA AI/ML-enabled device authorizations, 1995–2025.

## Discussion

We performed a cross-sectional analysis of all 1,430 AI/ML-enabled medical device authorizations recorded by the FDA between September 1995 and December 2025. Our analysis extends prior published reports of this regulatory record [1,2,3] by approximately two additional years and approximately 414 additional authorizations relative to the most recent published taxonomic analysis [3]. Three findings warrant discussion: the persistent concentration of authorizations within a small number of imaging-related specialties; the very small number of authorizations corresponding to several specialties with substantial clinical volume in the United States; and the bimodal distribution of authorizations across manufacturers.

The persistent dominance of radiology in the FDA AI/ML authorization record has been described in earlier analyses [1,2,3]. Our updated cumulative figure of 76.5% for the radiology share is consistent with reports from earlier time points. The stability of this share over a decade, despite a roughly 50-fold increase in the annual authorization rate, suggests that the underlying drivers are structural rather than transitional. Plausible structural drivers include the relative availability of large, labeled, standardized imaging datasets through Digital Imaging and Communications in Medicine (DICOM) infrastructure—the universal standard format and exchange protocol for medical imaging that enables interoperable storage and transfer of images across vendors and institutions, and that has historically facilitated assembly of training datasets at scale in radiology that are not as readily available in other specialties; the comparatively well-defined regulatory framework for image analysis software; and the existence of established commercial pathways for incorporating image analysis software into radiology workflows.

The corresponding underrepresentation of other clinical specialties is the more consequential finding. Several specialties that account for substantial volumes of clinical activity in the United States were associated with very small numbers of authorizations in the cumulative record. The absence of authorizations under a psychiatry or behavioral health review panel is particularly notable given the substantial population burden of mental illness and the active research literature on AI applications in mental health. Similarly, the small number of authorizations under the Pathology, Microbiology, and Obstetrics and Gynecology panels stands in contrast to the breadth of clinical activity in those specialties.

The manufacturer-level findings indicate a market structure in which a small number of repeat-authorizing firms account for a disproportionate share of authorizations, while most firms in the dataset have a single authorization. This pattern is consistent with both an emerging-industry interpretation, in which many small firms compete in early stages and a smaller number consolidate over time, and with a barriers-to-entry interpretation, in which the resources required for repeat regulatory engagement favor larger, established firms.

Public-facing aggregator resources that catalog the FDA list alongside additional metadata, including PhysicianAITools.com (https://physicianaitools.com), can support broader landscape analyses by linking the FDA’s authorization record with discovery-based information on medical AI tools that may not be captured by the FDA’s curated list.

### Limitations

This analysis has several limitations. First, the FDA’s published list of AI/ML-enabled devices is curated by the agency, and prior work has noted that the list is updated irregularly and does not include every device that may incorporate AI/ML functionality [3,5]. Second, the FDA’s lead review panel classification reflects regulatory categorization and may not align exactly with clinical specialty-level care delivery. Third, the dataset analyzed here did not contain information on indication for use, target patient population, performance characteristics, or post-market experience [7]. Fourth, the analysis was descriptive and did not include formal hypothesis testing. Finally, our manufacturer-concentration analysis treated each company name as a unique entity and did not account for corporate parent-subsidiary relationships.

## Conclusions

The cumulative FDA regulatory record of AI/ML-enabled medical devices through December 2025 contains 1,430 authorizations distributed unevenly across clinical specialties. The Radiology panel accounted for more than three-quarters of all authorizations, and the radiology share of new annual authorizations has remained stable across the past decade. Several specialties associated with substantial clinical activity in the United States are represented by very small absolute numbers of authorizations. The market is composed of a large number of single-authorization firms together with a small number of repeat-authorizing firms that account for a disproportionate share of authorizations. These findings are intended to inform discussion of where regulatory, infrastructure, and dataset investments could broaden the clinical scope of medical AI rather than to argue for any specific policy intervention.

## Disclosures

### Conflicts of interest

Pouyan Golshani, MD is the founder of GigHz (https://gighz.com), founded April 2025. GigHz operates the following platforms, all of which are disclosed in the interest of full transparency: GigHz Precision AI (radiology report assistant with embedded guideline-aware logic; https://gighz.com/radiology-report-assistant/); Pogosh CDS (clinical decision support API covering 413 conditions across 40+ specialties; https://gighz.com/pogosh/); CenterIQ (hospital price transparency database; https://gighz.com/centeriq/); PhysicianAITools.com (FDA AI/ML medical device catalog); Nakod (radiology prior authorization tool); GigHz Physician Finance Hub (tax planning and loan-decision tools for physicians; https://gighz.com/physician-finance/); LaVascular.com (consumer healthcare pricing); Repit.org (geographic and workforce data); Guide.MD (physician and hospital directory, in development). None of these platforms or their proprietary data were used as the primary data source for the present analysis, which relied solely on the FDA’s publicly available list of AI/ML-enabled medical devices. PhysicianAITools.com is mentioned in the Methods and Discussion sections as a public-facing aggregator that integrates the FDA list with additional metadata; this mention is not based on any analysis of PhysicianAITools’ proprietary metadata. Ms. Joseph reports no conflicts of interest.

### Funding

No external funding was received for this work.

### Author contributions

Pouyan Golshani, MD (ORCID 0000-0002-2042-4906) conceived the study, performed the data analysis, drafted the manuscript, and is the guarantor of the work. Mary Joseph, BS candidate (Undergraduate Researcher, Department of Biochemistry, University of California, Riverside, California, USA) performed independent data verification, conducted the literature review and reference verification, and contributed to manuscript revision. Both authors approved the final manuscript.

### Data availability

The FDA list of AI/ML-enabled medical devices is publicly available at https://www.fda.gov/medical-devices/software-medical-device-samd/artificial-intelligence-and-machine-learning-aiml-enabled-medical-devices.

## Notes

### Competing Interest Statement

Dr. Golshani is the founder of GigHz (https://gighz.com), founded April 2025. GigHz operates the following platforms, all of which are disclosed in the interest of full transparency: GigHz Precision AI (radiology report assistant; https://gighz.com/radiology-report-assistant/); Pogosh CDS (clinical decision support API; https://gighz.com/pogosh/); CenterIQ (hospital price transparency; https://gighz.com/centeriq/); PhysicianAITools.com (FDA AI/ML medical device catalog); Nakod (radiology prior authorization); GigHz Physician Finance Hub (https://gighz.com/physician-finance/); LaVascular.com (consumer healthcare pricing); Repit.org (geographic and workforce data); Guide.MD (physician/hospital directory, in development). None of these platforms or their proprietary data were used as the primary data source for the present analysis, which relied solely on the FDA's publicly available list of AI/ML-enabled medical devices. PhysicianAITools.com is mentioned in the Methods and Discussion sections as a public-facing aggregator that integrates the FDA list with additional metadata; this mention is not based on any analysis of PhysicianAITools' proprietary metadata. Ms. Joseph reports no conflicts of interest.

